# Clinical Study Applying Machine Learning to Detect a Rare Disease: Results and Lessons Learned

**DOI:** 10.1101/2021.12.07.21267403

**Authors:** William R. Hersh, Aaron M. Cohen, Michelle M. Nguyen, Katherine L. Bensching, Thomas G. Deloughery

## Abstract

Machine learning has the potential to improve identification of patients for appropriate diagnostic testing and treatment, including those who have rare diseases for which effective treatments are available, such as acute hepatic porphyria (AHP). We trained a machine learning model on 205,571 complete electronic health records from a single medical center based on 30 known cases to identify 22 patients with classic symptoms of AHP that had neither been diagnosed nor tested for AHP. We offered urine porphobilinogen testing to these patients via their clinicians. Of the 7 who agreed to testing, none were positive for AHP. We explore the reasons for this and provide lessons learned for further work evaluating machine learning to detect AHP and other rare diseases.

## Introduction

Machine learning has great potential to improve healthcare in many ways, from automated identification of patients eligible for testing and interventions to aid in diagnosis and risk stratification.(1,2) While predictive models to improve diagnosis and treatment have been developed and validated in many areas of medicine, relatively few models have been studied with real patients to determine their effectiveness in real-world settings.(3)

One application of machine learning is its use to identify patients who may have rare diseases. There are an estimated 1200 rare diseases that occur in humans.(4) These diseases often go undiagnosed for many years because they may resemble common diseases and their infrequent occurrence may result in physicians not considering them in diagnostic workups.(5) They may also have genetic components that may require specialized testing. Processing of data in the electronic health record (EHR) may help determine the presence of such diseases.(6) Although there are limitations to relying on EHR data for clinical research,(7) it was the only data available to us for these patients.

One rare disease for which effective treatment is available that may be amenable to EHR-based detection is acute hepatic porphyria (AHP). AHP is a subset of porphyria that refers to a family of rare, metabolic diseases characterized by potentially life-threatening acute attacks and, for some patients, chronic debilitating symptoms that negatively impact daily functioning and quality of life.(8) During attacks, patients typically present with multiple signs and symptoms due to dysfunction across the autonomic, central, and peripheral nervous systems and manifesting in abdominal pain, central nervous system abnormalities, and peripheral neuropathy. The prevalence of diagnosed symptomatic AHP patients is about 1 per 100,000.(9) Due to the nonspecific symptoms and the rare nature of the disease, AHP is often initially overlooked or misdiagnosed. One study demonstrated that diagnosis of AHP is delayed on average by up to 15 years.(10)

The predominant cause of AHP is a genetic mutation leading to a partial deficiency in the activity of one of the eight enzymes responsible for heme synthesis. AHP has low penetrance (~1%) and families carrying the gene may have few or only one affected member. For this reason, family history can be a poor diagnostic tool for this disease. The typical diagnostic approach for AHP is relatively inexpensive (typically around US$70) biochemical testing of random/spot urine for porphobilinogen.(11,12) Treatment of AHP has historically focused on avoidance of attack triggers, management of pain and other chronic symptoms, and treatment of acute attacks through the use of intravenous hemin. Recently, a new small interfering RNA (siRNA) molecule, givosiran, has been found to reduce the occurrence of acute attacks and impact other manifestations of the disease.(13)

We previously applied machine learning to an extract of 205,571 EHR records (200,000 selected from those receiving primary care at OHSU enriched with 5,571 patients with presence of the word “porph” in the diagnoses, procedures and notes of the record) from Oregon Health & Science University (OHSU) to determine whether this approach could be effective in identifying patients not previously tested for AHP, and who might be candidates to receive a diagnostic workup for AHP.(14) As described in the previous paper, we manually reviewed the records of 47 unique patients with the ICD-10-CM code E80.21 (Acute intermittent [hepatic] porphyria) and identified 30 who were true-positive cases. (The 30 cases were higher than would be expected in the general population likely due to our academic medical center is a tertiary referral center for Oregon and beyond.) We parsed the record into features, which were scored by frequency of appearance and filtered using univariate feature analysis. We manually choose features not directly tied to provider attributes or suspicion of the patient having AHP. Based on training with the rest of the dataset considered to be negative cases, the best cross-validation performance came from a support vector machine (SVM) algorithm using a radial basis function (RBF) kernel. The trained model was applied back to the full data set and patients were ranked by margin distance. The top 100 ranked negative cases were manually reviewed for symptom complexes similar to AHP, finding 22 patients who had no other diagnoses that could explain their symptoms. This study aimed to determine if our machine learning algorithm could identify patients with AHP in a clinical follow-up study.

## Methods

We obtained IRB approval to contact the 22 patients to offer free urine porphobilinogen testing (OHSU IRB00020617). Respecting the patient-clinician relationship, we structured the research protocol such that we would first obtain consent and permission to contact the patient from their primary care provider (PCP; physician, nurse practitioner, or physician assistant) or, when that the PCP could not be identified, the provider having the most clinical interaction with the patient in the OHSU record system. If approval was obtained by the provider, we would then contact the patient to obtain their consent and plan for participation. As this study was done during the COVID-19 pandemic, most interaction was done by telephone or email.

## Results

Of the 22 patients, six patients were either deceased or had moved from the Portland, Oregon area and had no follow-up address in the OHSU EHR, and we were unable to identify or contact their PCPs or other providers. Four other patients had moved their care to other local institutions and we were able to identify their PCPs. Eight patients had PCPs at OHSU, while four had non-PCP providers at OHSU significantly involved in their care.

Of the 16 patients for whom a clinician could be contacted, 13 agreed to allow us to contact their patient. (The rest did not believe testing would be clinically appropriate and declined to allow us to contract their patients.) We attempted to contact all 13 patients by phone and/or email. Four of these patients never replied to our attempts to contact them, with nine agreeing to sign the consent and come for testing. However, one patient never returned the consent form and another patient changed their mind about participation after signing the consent.

This resulted in seven patients being tested. All of these patients had virtually undetectable urine porphobilinogen, i.e., a normal test ruling out AHP.

**Figure 1.**
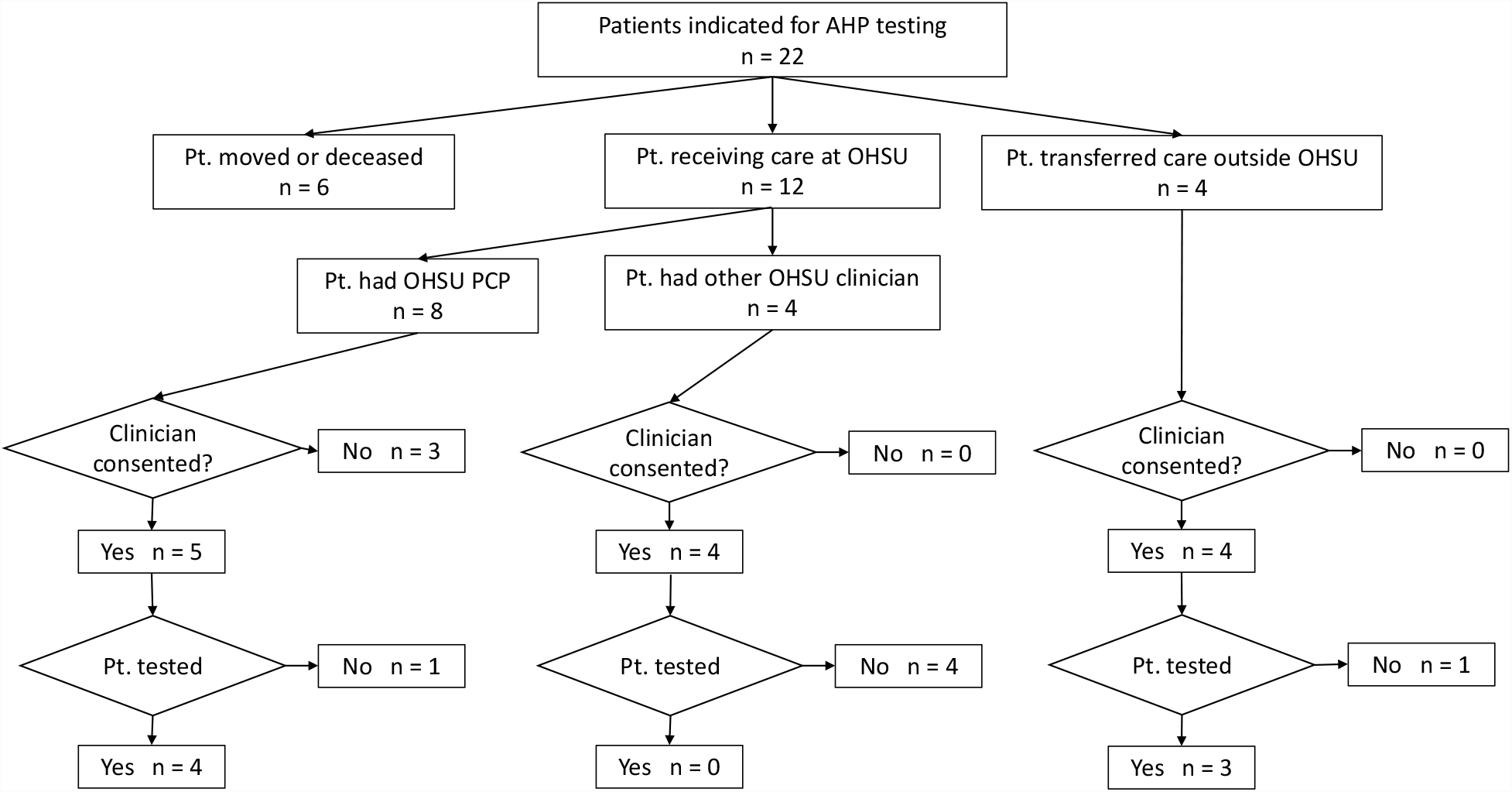
identification and testing of patients for attempted diagnosis of acute intermittent porphyria.

## Discussion

Clinical validation of machine learning is a critical step to its application in patient care. Being able to detect patients with rare diseases, whose diagnosis is often missed or delayed, could be an important use case, especially when effective treatments exist. We developed what seemed to be a promising approach to detecting AHP. All of patients identified by our algorithm had the classic neurovisceral symptoms of AHP, had no other diagnosis to explain those symptoms, and had never been tested for the disease.

The results of this study did not identify any patients with AHP confirmed by diagnostic testing. However, our algorithm did identify at least seven patients who had unexplained symptoms and that were appropriate candidates for diagnostic testing to rule out AHP, as confirmed by their PCP.

### Lessons Learned

This study both reiterates the need to validate machine learning approaches in medicine and highlights the challenges of doing so. In addition, there were lessons learned for the specific machine learning use case of rare disease detection.

#### Rare diseases detection is challenging to validate

By definition, the prevalence of a rare disease is very low. This will lead to a very low positive predictive value for most rare disease models, even if the lift (improvement factor of precision) is high. In our model, for example, cross-validation estimated the average precision to be 6%, which is quite low, but 400 times what would be expected from random sampling (0.015%). The disease is rare enough that it was unlikely that we would identify a *confirmed* case of previously undiagnosed AHP, even with a 400-factor improvement in detection.

The small number of positive samples available for learning and model creation results in a lack of positive samples for evaluation. Standard methods of cross-validation and training/testing data set splitting are difficult to use meaningfully with small numbers of positive samples. This is likely to be a continuing challenge for applying ML to rare diseases and new modeling and evaluation approaches will be needed. Here, we used an approach inspired by information retrieval research, using the model to essentially score similarly with the known positives and conducted extensive manual review and clinical follow up on the predictions in order to perform the evaluation.

Furthermore, EHR data is known to be noisy and incomplete, resulting in further challenges. These limitations must be acknowledged for any automated approach trying to identify undiagnosed cases of rare disease. Integration of other data sources or available of a “complete” medical record may help improve this.

#### Learning to work together

Another lesson learned in this study concerned the challenges of machine learning researchers who are not part of the care teams of patients. This necessitated the two-step approach of first contacting the PCP and obtaining approval to contact the patient. Another challenge was our looking for a rare disease with which most PCPs had little experience.

Our study was also likely hindered by the complex recruitment process that was necessitated by respecting the patient-clinician relationship. Several steps were required to reach busy clinicians to obtain their consent and participation, which then required follow-on with patients who could be difficult to contact. Finally, the fact that we attempted to carry out this study during the COVID-19 pandemic made everything more difficult due to patients’ hesitancy to visit healthcare institutions.

#### Even Rarer When Looking for Undiagnosed Patients

Even for those patients who were tested, there were a number of reasons why they had negative results. To begin with, AHP is a rare disease that was already over-represented relative to its population incidence at our tertiary medical center. As such, there may be few patients for whom to make a diagnosis *de novo*.

Another possibility is that our algorithm selected the wrong patients for testing, i.e., there may be patients who receive care at our medical center but did not rank high enough in the output of our model to be assessed for the need for testing. However, for the patients tested their PCP did confirm that testing was appropriate, so possibly our algorithm cut off point should be extended in the future to include more patients.

An additional challenge for this study is the nature of the symptoms of patients who have AHP. Most of these are non-specific yet can be debilitating to patients, often leading to years of frequent clinical visits and substantial use of diagnostic testing, including expensive imaging procedures.

#### In some rare diseases, diagnostic testing may be expensive

One of the hopes for detection of AHP is the relatively modest expense of diagnosis, consisting of an elevated random/spot urine porphobilinogen.(12) This is unlikely to be the case for many rare diseases where more expensive and invasive diagnostic testing might be required. Thus, the approach of aiming to detect rare diseases must be balanced with the added cost and possible risk to patients, especially if the probability of such detection is low.

## Conclusion

Despite our lack of success in identifying any new AHP patients, the study did show value in identifying symptomatic patients where a diagnostic test was appropriate (confirmed by chart review and PCP) and had not yet been done, perhaps improving the completeness of their care. We hope that our experience in trying to conduct this study will inform the community and encourage other researchers to attempt to clinically validate their machine learning systems to help move these approaches from the basic science towards improving clinical care.

## Data Availability

Not available.

